# Mitral Annular Disjunction in Heritable Thoracic Aortic Disease: Insights From the Montalcino Aortic Consortium

**DOI:** 10.1101/2024.05.06.24306968

**Authors:** Kishan L. Asokan, Jennifer R. Landes, Wannes Renders, Laura Muiño Mosquera, Julie De Backer, David W Jantzen, Anji T. Yetman, Gisela Teixido-Tura, Arturo Evangelista, Richmond Jeremy, Edward G. Jones, Shaine Morris, Tam Doan, Maral Ouzonian, Alan Braverman, Guillaume Jondeau, Olivier Milleron, Siddharth K. Prakash, Dianna M. Milewicz

**Author notes:** Corresponding Author: Siddharth K. Prakash, MD, PhD Department of Internal Medicine John P and Kathrine G McGovern Medical School University of Texas Health Science Center at Houston 6431 Fannin Street MSB 6.116 Houston, Texas 77030 USA.

## Abstract

**Background:** Mitral annular disjunction (MAD), posterior displacement of the mitral valve leaflet hinge point, predisposes to arrhythmias or sudden cardiac death. We evaluated the burden of MAD, mitral valve prolapse (MVP), and mitral regurgitation (MR) by heritable thoracic aortic disease (HTAD) gene in a cross-sectional analysis of 2014-2023 data in the Montalcino Aortic Consortium (MAC) registry.

**Methods:** MAD was determined by direct measurement of echocardiographic images. MR and MVP were defined according to current clinical guidelines. Associations were evaluated using chi-squared or Fisher exact tests.

**Results:** MR and MVP were enriched in MAC participants (672) with pathogenic variants (PV) in TGF-β pathway genes. The combination of MR and MVP was associated with mitral surgery and arrhythmias. In the subgroup with available images, MAD was enriched in *SMAD3* PV compared to other TGF-β PV (PR 1.8 [1.1-2.8], *P*< 0.02). Severe disjunction (>10 mm) was only observed in the TGF-β subgroup and was further enriched in participants with *SMAD3* PV (PR 3.1 [1.1-8.6]). MVP (PR 5.2 [3.0-9.0]) and MR (PR 2.7 [1.8-3.9) were increased in participants with MAD, but MAD was not independently associated with adverse cardiac or aortic events.

**Conclusions:** Mitral phenotypes are more prevalent in individuals with PV in TGF-β pathway genes, particularly *SMAD3*, and are associated with adverse aortic and cardiac events. Because congenital mitral disease may be the primary presenting feature of SMAD3 PV, genetic testing for HTAD should be considered for such individuals, especially if they also have a family history of HTAD.

**Clinical/Research Perspective:** *What Is New?:*

1) Mitral regurgitation, mitral valve prolapse, and mitral annular disjunction (MAD) are common in heritable thoracic aortic disease (HTAD) caused by pathogenic variants (PV) in TGF-β pathway genes (*SMAD3*, *TGFBR1*, *TGFBR2*, *TGFB2*, or *TGFB3*) and are associated with adverse cardiac events.
2) Pathological mitral phenotypes are particularly prominent in people with *SMAD3* PV and may be the presenting feature of HTAD in some cases.

*What Are the Clinical Implications?:*

3) Pathological mitral valve phenotypes may identify a high-risk subgroup of HTAD cases with more frequent adverse cardiovascular events. Because congenital mitral disease may be the primary presenting feature of *SMAD3* PV, genetic testing for HTAD should be considered for such individuals, especially if they also have a family history of HTAD.

## Introduction

Mitral annular disjunction (MAD) is atrial displacement of the mitral valve leaflet hinge point from the ventricular myocardium. MAD was first described in autopsy studies of individuals with mitral valve prolapse (MVP) who experienced sudden cardiac death (SCD) [1, 2]. MVP or MAD may pose an increased risk for ventricular arrhythmias and SCD, and MAD is an independent risk factor for arrhythmic events [2–8]. Cardiac magnetic resonance studies identified myocardial fibrosis in the inferolateral left ventricle of MAD cases and implicated significant fibrosis as a nidus for ventricular arrhythmias [6]. In Marfan syndrome (MFS), MAD also predisposes to ventricular tachycardia and SCD, and displacement of the mitral hinge point > 10 mm is correlated with the likelihood of ventricular arrhythmias [4, 9]. The genetic cause of most sporadic MAD cases and potential associations between pathogenic variants (PV) in other HTAD genes and MAD are unknown.

The Montalcino Aortic Consortium (MAC, NCT04005976) is an international collaboration of expert HTAD clinical centers that was established in 2013 to investigate the disease course of individuals with validated PV in a clinically actionable HTAD gene [10, 11]. Genetic syndromes (e.g. Marfan syndrome or Loeys-Dietz syndrome) cause approximately 5% of HTAD cases, with the vast majority classified as non-syndromic [12]. Known causative HTAD genes encode extracellular matrix proteins (*FBN1*, *COL3A1*, *LOX*), components of the TGF-β signaling pathway (*SMAD2*, *SMAD3*, *SMAD4*, *TGFB2*, *TGFB3*, *TGFBR1*, *TGFBR2*), and smooth muscle cell contractile components (*ACTA2*, *MYLK*, *PRKG1*, *MYH11*). [12,13][16]. More than 20% of individuals with non-syndromic HTAD have a family history of HTAD, signaling the urgent need for early diagnosis and surveillance of affected individuals [13, 14]. The MAC data on the natural history of HTAD due to specific PV have informed new clinical recommendations for management of HTAD based on the underlying mutated gene in the 2022 ACC/AHA Guidelines for the Diagnosis and Management of Aortic Disease [13, 17]. The specific type of PV may influence the timing of elective surgery to prevent an aortic dissection because some PV are associated with dissections at smaller absolute aortic diameters.

Aneurysms and dissections of smaller arteries and congenital cardiac malformations such as bicuspid aortic valve and septal defects are more common in some HTAD PV carriers [18][19][20]. We hypothesize that mitral valve prolapse (MVP), mitral regurgitation (MR), or MAD may be enriched in groups with HTAD PV due to phenotypic overlap between MAD, MFS, and HTAD. This study focused on the analysis of mitral phenotypes including MAD in the MAC cohort and potential associations with the mutated HTAD gene.

## Methods

We conducted a multicenter cross-sectional study of MAC registry participants. The study protocol was reviewed and approved by the Committee for the Protection of Human Subjects at the University of Texas Health Science Center at Houston (IRB HSC-MS-16-0191). The investigators accessed person-level data from a single registry to create the study population. Additional data or clarifications were requested from the enrolling site investigators as needed. The data collection period was from 12/2014 to 8/2023. We considered pathogenic and likely pathogenic variants as a single group for analysis (referred to as PV). Participants without validated HTAD gene PV were excluded from the analysis. We also excluded individuals with *FBN1* PV because MAC exclusively enrolled pediatric *FBN1* cases at the time of this analysis.

The primary independent variable was mitral disease, defined as any degree of mitral valve prolapse (MVP), or mild, moderate, or severe mitral regurgitation (MR). The primary dependent variables were clinical outcomes: the diagnosis of any arrhythmia, congestive heart failure (CHF), or aortic dissection, or aortic or mitral valve surgery. The principal covariates were sex and the HTAD PV. For the subgroup of MAC participants with available images, the primary independent variable was MAD.

MAD distance was measured by direct review of echocardiogram images. We determined that MAD was present if we measured at least three millimeters between the posterior mitral valve leaflet hinge point and the inferolateral myocardium using the inner edge to inner edge method [5]. MAD was measured during mid-systole at the peak of the T wave in the parasternal long axis view. Two experienced reviewers reported the mean of three measurements for each cine series.

Parasternal long axis images were rated using American Society of Echocardiography criteria for orientation and image quality. Orientation was acceptable if the horizontal septum deviated less than 30 degrees from the horizontal plane, the proximal aorta aortic and mitral valves were visible, and the ventricular septum was visible to the apex. Image quality was assessed by brightness level, resolution, and focus on the region of interest. Reviewers rated each category as optimal, adequate, or inadequate. If any category was inadequate, the image was excluded from analysis.

Pearson’s chi-squared tests or Fisher exact tests were used for comparisons of categorical data. Quantitative data were described using median values with interquartile ranges. The strength of associations was assessed by calculating the E-values of prevalence ratios (PR) [21]. A multivariate regression model was constructed to evaluate associations between age, sex, mutation type, MAD, MVP, and MR and the composite outcome of aortic surgery, mitral surgery, aortic dissection, arrhythmia, or CHF using DATAtab (DATAtab e.U. Graz, Austria).

## Results

From an initial sample of 1080 MAC participants, 130 subjects were excluded due to missing genotypes. 203 participants were excluded due to missing MR or MVP phenotypes, and 75 were excluded for missing variant pathogenicity classifications. A total of 672 MAC participants (51% male, median age 36 years [IQR 19,51]) were included in this study: 312 (46%) had PV in TGF-β pathway genes. Images from 240 participants were used to assess MAD (50% male, median age 33 years [IQR 20,33]). Nine subjects were excluded from the MAD sub-study due to image quality. Three quarters of MAC participants had a PV in an HTAD gene (Table 1). Participants with variants of uncertain significance (n=155) were excluded from this analysis.

**Table 1.** Characteristics of Study Cohort (n=672)

| Characteristic | Value | Missing |
| --- | --- | --- |
| Female (%) | 329 (49) | 0 |
| Height, cm (IQR) | 173 (163,183) | 64 |
| Weight, Kg (IQR) | 74 (58,93) | 57 |
| Average age at enrollment, y (IQR) | 36 (19,51) | 4 |
| Mitral valve prolapse (%) | 99 (15) | 0 |
| Mitral regurgitation (%) | 149 (22) | 0 |
| Mitral images available (%) | 240 (36) | 432 |
| Pathogenic or likely pathogenic variants (%) | 517 (77) | 0 |
| Variants of uncertain significance (%) | 155 (23) | 0 |
| TGF- $\beta$ pathway variants (%) | 443 (66) | 0 |

cm: centimeters, kg: kilograms, y: years, IQR: interquartile range; Missing: number of MAC participants with missing data.

### Mitral Valve Disease in the MAC Cohort

The prevalence of MR and MVP in the United States Population is estimated at 2-3% [23, 24]. In the MAC cohort, the overall prevalence of MR was 22%, the prevalence of moderate or severe MR was 3%, and the prevalence of MVP was 14% (Table 1). Therefore, MR and MVP are 5-10 times more prevalent in MAC than in the general population. There were no significant differences in the prevalence of MR or MVP in males vs. females, or in individuals with PV compared to individuals with VUS (Figure S1 and Figure S2).

Most individuals with MVP had PV in TGF-β pathway genes (Table 2). Individuals with *SMAD3*, *TGFBR1*, *TGFBR2*, *TGFB2*, or *TGFB3* PV comprised 60% of the MAC cohort with PV but 90% of MVP and 71% of MR cases (Figure S3). The number of MAC participants with *ACTA2* and *TGFBR2* PV were similar, but MVP was seven times more common in the subgroup with *TGFBR2* PV compared to the subgroup with *ACTA2* PV (17/83 (20%) vs. 3/89 (3%), PR 6.1 [1.8-20], *P*<0.001).

**Table 2.**
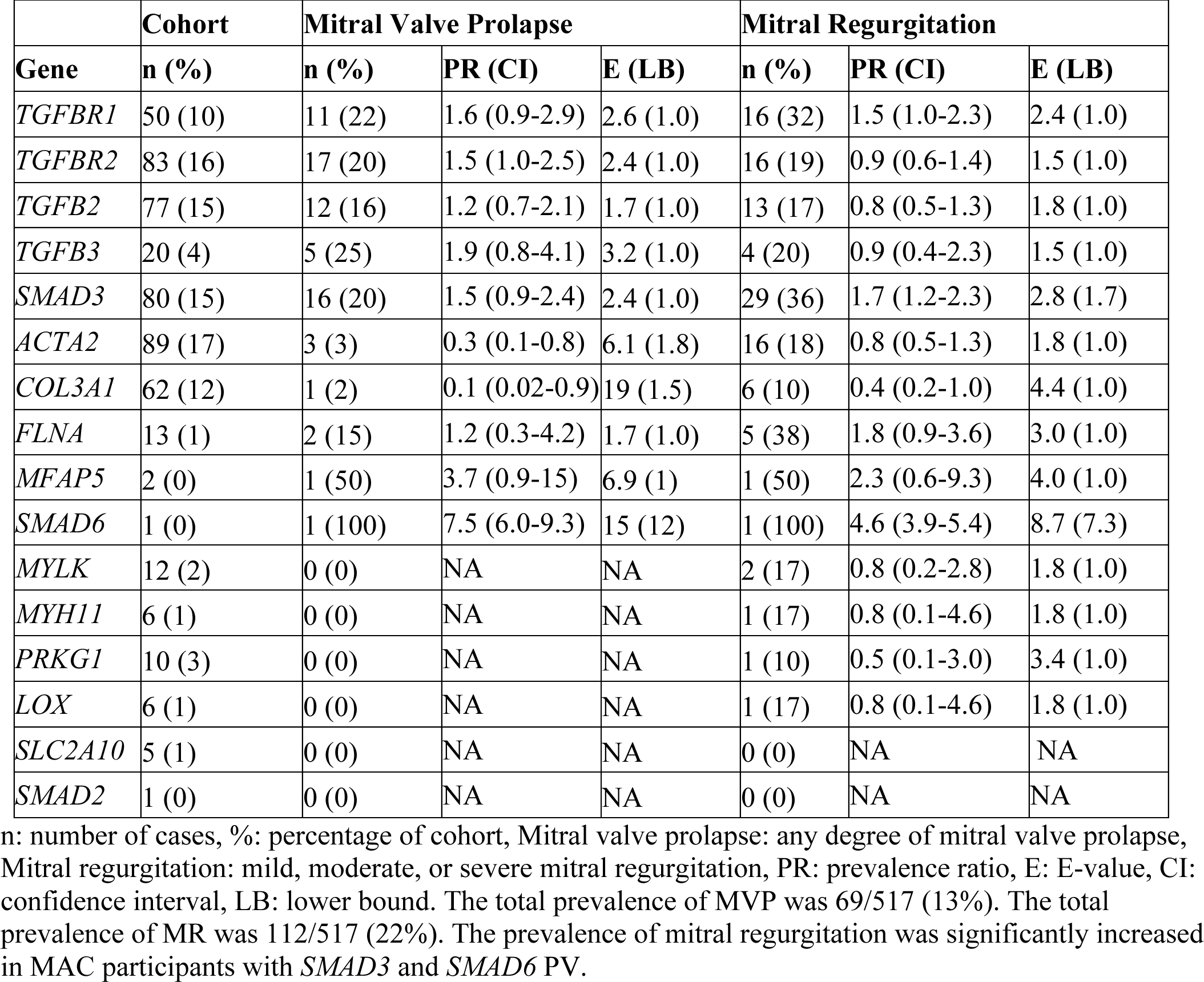
Mitral Valve Prolapse and Mitral Regurgitation by Gene.

The proportion of individuals with MVP or mild, moderate, or severe MR was significantly enriched in *TGFBR1* and *SMAD3* PV carriers (Figure S3 and Figure S4). The prevalence of MR was more evenly distributed between groups with PV in other HTAD genes such as *ACTA2* (18%, Table 2).

Cardiac outcomes of MAC participants with mitral disease were significantly worse than the outcomes of participants who did not have MVP or MR. Mitral valve surgery (4/32 (13%) vs. 2/367 (0.5%), PR 23 [4.4-120], P<0.001) and arrhythmias (6/32 (19%) vs. 13/367 (4%), PR 5.3 [2.2-13], P<0.01) were significantly increased in the group of MAC participants with MVP and MR when compared to the group without MVP or MR (Table 3).

**Table 3.** Cardiac and Aortic Events by Mitral Phenotype.

| Events | No MR or MVP (%) | MR and MVP (%) | n (%) | PR (CI) | E (LB) |
| --- | --- | --- | --- | --- | --- |
| Aortic surgery | 105 (29) | 14 (44) | 175 (34) | 1.5 (1.0-2.3) | 2.4 (1.0) |
| Aortic dissection | 88 (24) | 9 (28) | 145 (28) | 1.2 (0.7-2.1) | 1.7 (1.0) |
| Mitral valve surgery | 2 (0.5) | 4 (13) | 12 (2) | 23 (4.4-120) | 45 (8.3) |
| CHF | 10 (3) | 1 (3) | 22 (4) | 1.1 (0.2-8.9) | 1.4 (1.0) |
| Arrhythmia | 13 (4) | 6 (19) | 30 (6) | 5.3 (2.1-13) | 10 (3.6) |

MR: mild, moderate, or severe mitral regurgitation, MVP: any degree of mitral valve prolapse, CHF: congestive heart failure, PR: prevalence ratio, E: E-value, CI: 95% confidence interval, LB: lower bound Percentages are in parentheses. Two-tailed *P*-values were calculated using Fischer's exact test. See Table S1 for complete outcomes data. Mitral valve surgery and arrhythmia were increased in MAC participants with MR and MVP.

The prevalence of moderate or severe MR was not significantly different in MAC participants with PV in TGF-β pathway genes compared to PV in other HTAD genes (*P*<0.06). Individuals with moderate or severe MR more frequently had aortic surgery (*P*<0.01), mitral surgery (*P*< 0.001), CHF (*P*< 0.001), and arrhythmias (*P*<0.01, Table S2). In the subgroup of participants with available images, the composite of MAD, MVP, and moderate or severe MR was more prevalent in participants with *SMAD3* PV compared to those with PV in other HTAD genes (34/67 (51%) vs. 41/172 (24%), PR 2.1 [1.5-3.0], *P*<0.001).

The proportions of probands and relatives who had MVP or MAD were not significantly different, but more probands exhibited the composite clinical endpoint. These findings suggest that the presentation of mitral valve disease was similar in probands and relatives (Table S3).

### Mitral annular disjunction in the MAC cohort

Most mitral images were from individuals with PV in TGF-β pathway genes (68%), consistent with the proportion of the TGF-β subgroup in the entire MAC cohort. MAD was observed in 24% (57/240) of the subgroup with available images (Table 4). MAD was more prevalent in MAC participants with PV in TGF-β pathway genes compared to participants with other PV (52/162 (32%) vs. 5/78 (6%), PR 5.0 [2.1-12], *P*<0.001, Figure 1). MAD was highly enriched in MAC participants with *SMAD3* PV compared to PV in other TGF-β pathway genes (29/67 (43%) vs. 23/95 (24%), PR 1.8 [1.1-2.8], P<0.02). MAD, MVP, and/or MR was more prevalent in participants with SMAD3 PV compared to participants with PV in other TGF-β genes (42/67 (63%) vs. 56/95 (41%), PR 1.5 [1.1-2.1], P<0.02). MAD was also enriched in MAC participants with PV in *TGFBR1*, *TGFBR2*, and *TGFB2* (Figure S5). The overall prevalence of MAD was not significantly different in males (23%, 27/120) compared to females (25%, 30/118, *P*<0.7) or in probands compared to relatives (Table S3).

**Figure 1.**
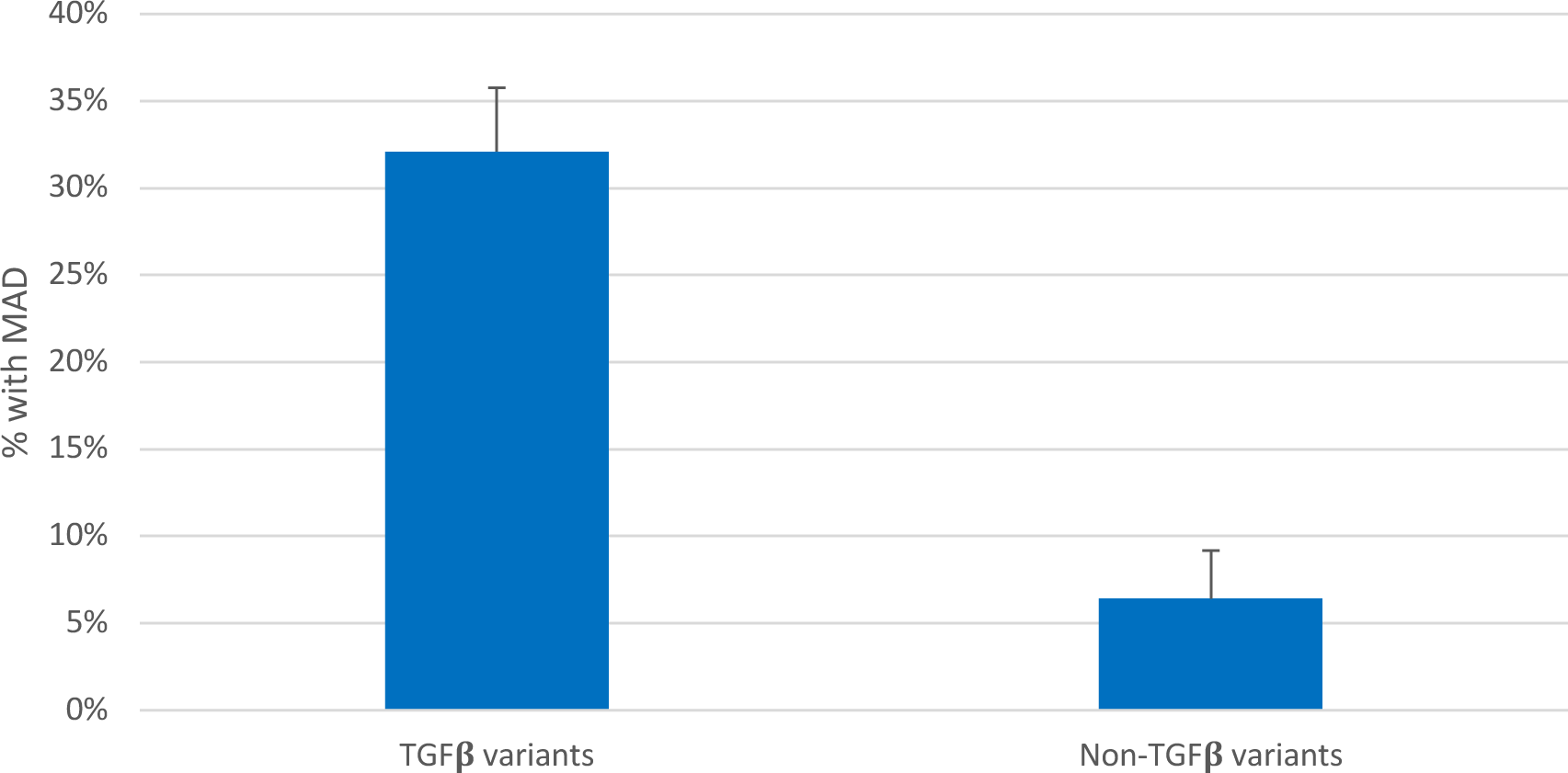
Prevalence of MAD in MAC participants with PV in TGF-β pathway genes compared to other HTAD genes. MAD: Mitral annular disjunction. TGF-β pathway genes are *SKI*, *SMAD3*, *SMAD6, TGFBR1*, *TGFBR2*, *TGFB2*, and *TGFB3*.

**Table 4.** Prevalence of MAD by Gene in MAC Participants with Available Images.

| **Gene** | **n (%)** | **MAD (%)** |
| --- | --- | --- |
| *TGFBR1* | 22 (9) | 6 (27) |
| *TGFBR2* | 45 (19) | 12 (27) |
| *TGFB2* | 22 (9) | 5 (23) |
| *TGFB3* | 5 (2) | 0 (0) |
| *SMAD3* | 67 (28) | 29 (43) |
| *SMAD6* | 1 (0) | 0 (0) |
| *ACTA2* | 29 (12) | 3 (10) |
| *COL3A1* | 26 (11) | 0 (0) |
| *LOX* | 5 (2) | 0 (0) |
| *MFAP5* | 2 (1) | 1 (50) |
| *MYH11* | 3 (1) | 1 (33) |
| *MYLK* | 1 (0) | 0 (0) |
| *PRKG1* | 2 (1) | 0 (0) |
| *FLNA* | 8 (3) | 0 (0) |
| *SCL2A10* | 2 (1) | 0 (0) |
| TGF-β PV | 162 (68) | 52 (32) |
| Other PV | 78 (32) | 5 (6) |
| Total | 240 | 57 (24) |

n: number of cases, %: percentage of cohort. TGF-β : *TGFBR1*, *TGFBR2*, *TGFB2*, *TGFB3*, *SMAD3*, *SMAD6*; MAD: Mitral annular disjunction.

The prevalence of MVP (26/57 (46%) vs. 16/183 (9%), PR 5.2 [3.0-9.0], *P*<0.001) and any degree of MR (30/57 (53%) vs 36/182 (20%), PR 2.7 [1.8-3.9], *P*<0.001), but not moderate or severe MR (8/183 (4%) vs. 4/57 (7%), *P*<0.3), was increased in MAC participants who had MAD. However, in a logistic regression model with age, sex, MR, MVP, and PV type, MAD was not independently associated with arrhythmia, aortic surgery, aortic dissection, or CHF in the entire MAC cohort or in participants with *SMAD3* PV (Figure S6). Left ventricular ejection fraction, left atrial volume, and the maximum diameter of the thoracic aorta also did not vary significantly based on the presence of MAD (Table 5).

**Table 5.**
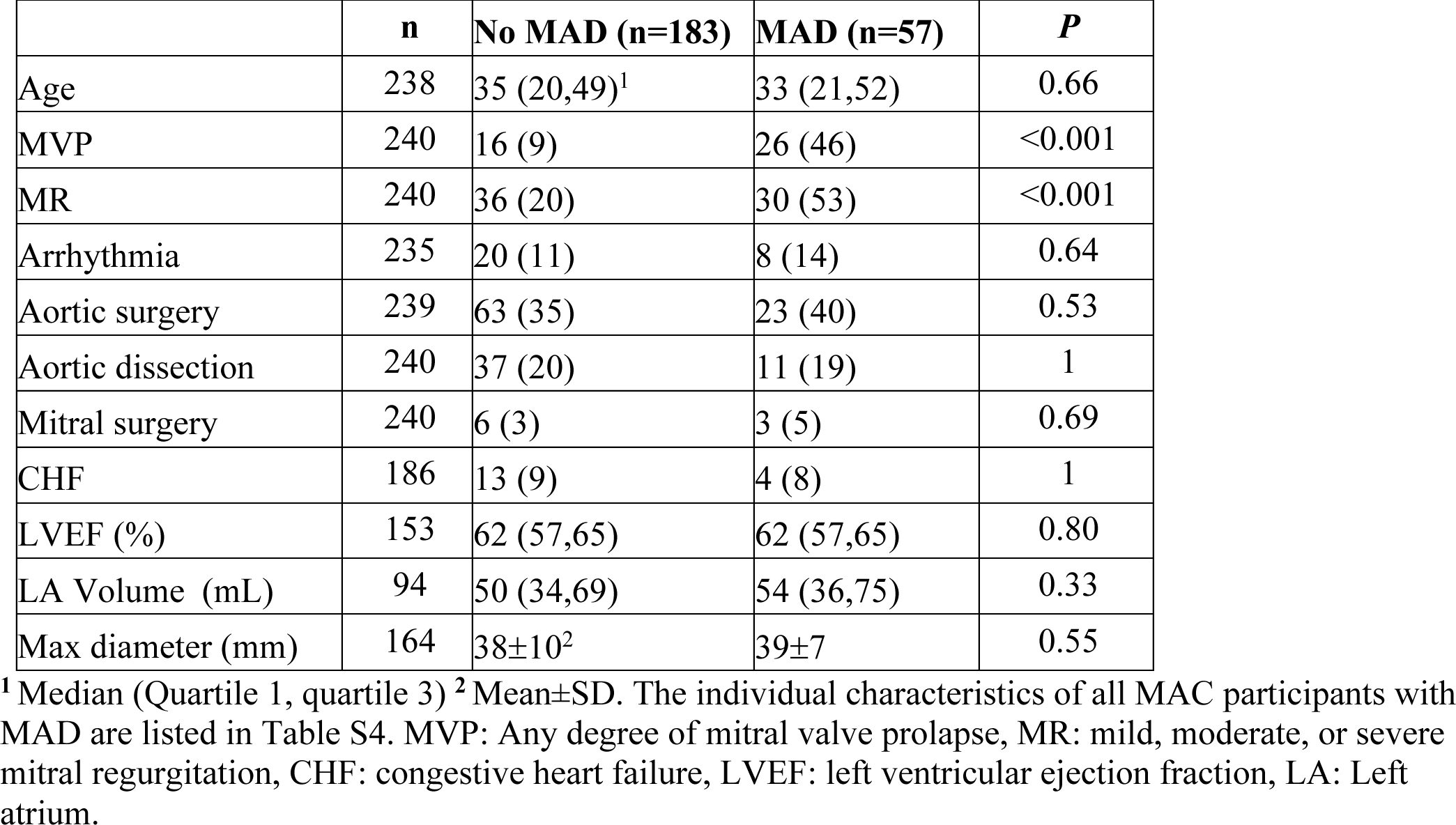
Characteristics of MAC Participants by MAD Phenotype.

The distance between the annulus and myocardial border was measured in 56 MAC participants, who were categorized as having mild (3-10 mm, 39) or severe (≥10 mm, 16) disjunction. Severe disjunction was only observed in MAC participants with TGF-β gene PV and was particularly enriched in the subgroup with *SMAD3* PV (11/67 (16%) vs. 5/95 (5%), PR 3.1 [1.1-8.6], *P*<0.03). Pathological mitral phenotypes (>mild MR, severe disjunction, or MAD with MVP) without significant aortic dilation (Z<3) were present only in MAC participants with PV in TGF-β pathway genes, were enriched in the subgroup with *SMAD3* PV (12/67 (18%) vs. 3/95 (3%), PR 5.7 [1.7-19], *P*<0.01, Table S4), and were significantly more prevalent in relatives compared to probands (Table S3). There was no significant difference in the prevalence of severe annular disjunction between males and females (7/120 (6%) vs. 9/118 (8%)).

## Discussion

In MAC participants with HTAD, we found that the prevalence of MVP was five times higher than in the general population and was frequently accompanied by MAD [23, 24]. Our data is consistent with other studies showing that PV in TGF-β pathway genes, primarily *TGFBR1* (22%, 11/50), *TGFBR2* (20%, 17/83), *TGFB3* (25%, 5/20), and *SMAD3* (20%, 16/80, are most strongly associated with these mitral phenotypes [4,25,26,31]. The overall rates of MVP and MAD were 7-8 times higher in MAC participants with PV in TGF-β pathway genes than in the rest of the MAC cohort. TGF-β dysregulation can increase matrix metalloproteinase activity and activate endothelial to mesenchymal transition in valve interstitial cells, predisposing to myxomatous mitral valve degeneration [27]. We also identified MAD in MAC participants who did not have PV in TGF-β pathway genes, primarily the subgroup with *ACTA2* PV [13]. These data show how the pathophysiology caused by PV in HTAD genes may extend beyond the aorta and highlight potential implications for PV carriers who are at increased risk for cardiac complications related to MVP, such as arrhythmias or CHF.

We observed that mitral phenotypes in MAC are significantly associated with aortic surgery, mitral surgery, and arrhythmias. It is unclear if mitral and aortic disease are causally linked, or if pathological mitral disease with MAD may reflect more severe connective tissue degeneration that predisposes to HTAD or myocardial fibrosis driving arrhythmias [28, 29, 31, 32]. The subgroup of MAC participants with MVP and MR appear to be at particularly high risk for adverse cardiac events. Our data indicate that this group may benefit from more intensive surveillance or targeted interventions [29, 30].

In contrast to MAD in MFS, we found that MAD was not independently associated with aortic or cardiac events in MAC participants. This is likely because most MAC participants with MAD also presented with MVP. In MFS, the arrhythmic risk that was attributed to MAD occurred predominately in the context of other mitral phenotypes, with arrhythmias detectable in 40% of individuals who had MVP and MAD compared to 20% of individuals with MVP and no MAD [8, 33]. In this study, the rate of arrhythmias in MAC participants who had both MAD and MVP (4/26, 15%) was similar to individuals who had MAD but no MVP (4/31, 13%). However, these observations do confirm that the risk for arrhythmias is markedly increased in the subgroup of individuals with HTAD who have pathological mitral phenotypes and suggest that ambulatory electrocardiographic screening may be beneficial.

Biases due to possible misclassification of individuals with mild mitral phenotypes and missing phenotypic data may have caused the effect sizes of associations between mitral and clinical phenotypes to be underestimated. Conversely, we acknowledge that the strength of some associations may be inflated by unmeasured confounders because a relatively limited set of phenotypes were available for study participants. In some cases, combined confounding from these factors could reach the magnitude of the E-value and could nullify a significant association. MAC study sites are referral centers that tend to recruit participants with more severe HTAD phenotypes than in community settings. Therefore, larger prospective studies are needed to corroborate these findings and to determine the prognostic significance of MAD in different genetic and clinical contexts.

## Data Availability

The authors declare that all supporting data are available within the article and its online supplementary files

## Acknowledgements

We thank the study participants and referring clinicians without whom this study would not have been possible. Special thanks to Kristin Ankoma-Sey and Walter Velasco-Torrez for assistance with MAC registry data. The Montalcino Aortic Consortium investigators are: Dianna M. Milewicz, MD, PhD; Reed Pyeritz, MD, PhD; Julie De Backer, MD, PhD; Catherine Boileau, PhD; Guillaume Jondeau, MD, PhD; Alan Braverman, MD; Shaine A. Morris, MD; Arturo Evangelista, MD; Maral Ouzounian, MD, PhD; Anji Yetman, MD; Richmond, Jeremy, MB, BS, PhD; Sherene Shalhub, MD, MPH; Eloisa Arbustini, MD; Lesley Ades, MD; Nanette Alvarez, MD; Anne Child MD; Bo Carlberg, MD, PhD; Ismail El-Hamamsy, MD, PhD; David Chitayat, MD; Anne De Paepe, MD, PhD; Richard B. Devereaux, MD; Bridgete Fernandez, MD; Josephine Grima, PhD; Maarten Groenink, MD, PhD; Gabrielle Horne, MD, PhD; Yskert von Kodolitsch, MD; Ronald Lacro, MD; Scott A. LeMaire, MD; Alex V. Levin, MD; David Liang, MD, PhD; Irene Maumenee, MD; Vivienne McConnell; Rocio Moran, MD; Hiroko Morisaki, MD, PhD; Takayuki Morisaki, MD, PhD; Alex Pitcher, PhD; Nancy Poirier, MD; Francesco Ramirez, PhD; Peter Robinson, MD; Dieter Reinhardt, PhD; Meike Rybczynski, MD; George Sandor, MD; Lynn Sakai, PhD; Denver Sallee, MD; Michael N. Singh, MD; Bert Callewaert, MD, PhD; Ellen Regalado, CGC; Marion A. Hofmann-Bowman, MD, PhD; Fabien Labombarda, MD; Laurence Faivre, MD, PhD; Claire Bouleti, MD, PhD; Marjolijn Renard, PhD; Derek Human, BA, BM. BCh; Zoltan Szabolcs, MD, PhD; Andrew Michael Crean, MD; Joseph Bavaria, MD; Apostolos Psychogios, MD; Vidyasagar Kalahasti, MD; Philip Giampietro, MD, PhD; Laura Muiño-Mosquera, MD; Gisela Teixido-Tura, MD, PhD; Olivier Milleron, MD; Mark Evan Lindsay, MD, PhD; Juan Manuel Bowen, MD; Shuping Ge, MD; Anthony Caffarelli, MD; Jolien Roos-Hesselink, MD; Mary B. Sheppard, MD; Andrew Choong, MBBS; Enid Neptune, MD; Zhou Zhou, MD, PhD; Alessandro Pini, MD; Germano Melissano, MD; Elisabetta M. Mariucci, MD; Elena Cervi, MD, PhD; Melissa L. Russo, MD; Pascal-Nicoloas Bernatchez, PhD; Mitra Esfandiarei, PhD; Julien Marcadier, MD; Seda Tierney, MD; Jessica Wang, MD, PhD; Kevin Michael Harris, MD; Sara B. MacKay, CCGC; Andrea L. Rideout, CCGC; Sandhya Prakash, MD; Anthony Martin Vandersteen, PhD; Nadine Hanna, PharmD, PhD; Carine Le Goff, PhD; Quentin Pellenc, MD; Patrick Sips, PhD; Scott Michael Damrauer, MD; Lois Starr, MD, PhD; Bo Yang, MD, PhD; Jay B. Shah, MD; Justin T Tretter, MD; Irman Forghani, MD; Kelly Cox, MD; Siddharth K. Prakash, MD, PhD; Sachiko Kanki, MD, PhD; Justin A Weigand, MD; Taylor A. Beecroft, CGC; Alan Frederick Rope, MD; Yaso Emmanuel, MBChB, DPhil.; David Dichek, MD; Michelle Keir, MD; Klaus Kallenbach, MD; Talha Niaz, MBBS; Mirela Tuzovic, MD; Anthony L. Estrera, MD; Timothy Carter, MD; Martin Czerny, MD; Anna Sabaté Rotés, MD, PhD; Brett Carroll, MD; William T. Brinkman, MD; David R. Murdock, MD; Christopher Phillips Jordan, MD.

## Nonstandard Abbreviations and Acronyms

LDS: Loeys-Dietz syndrome
MAD: Mitral Annular Disjunction
MFS: Marfan syndrome
SCD: Sudden Cardiac Death
HTAD: Heritable Thoracic Aortic Disease
TGF-β: Transforming Growth Factor-Beta

## Supplemental Material

**Figure S1.**
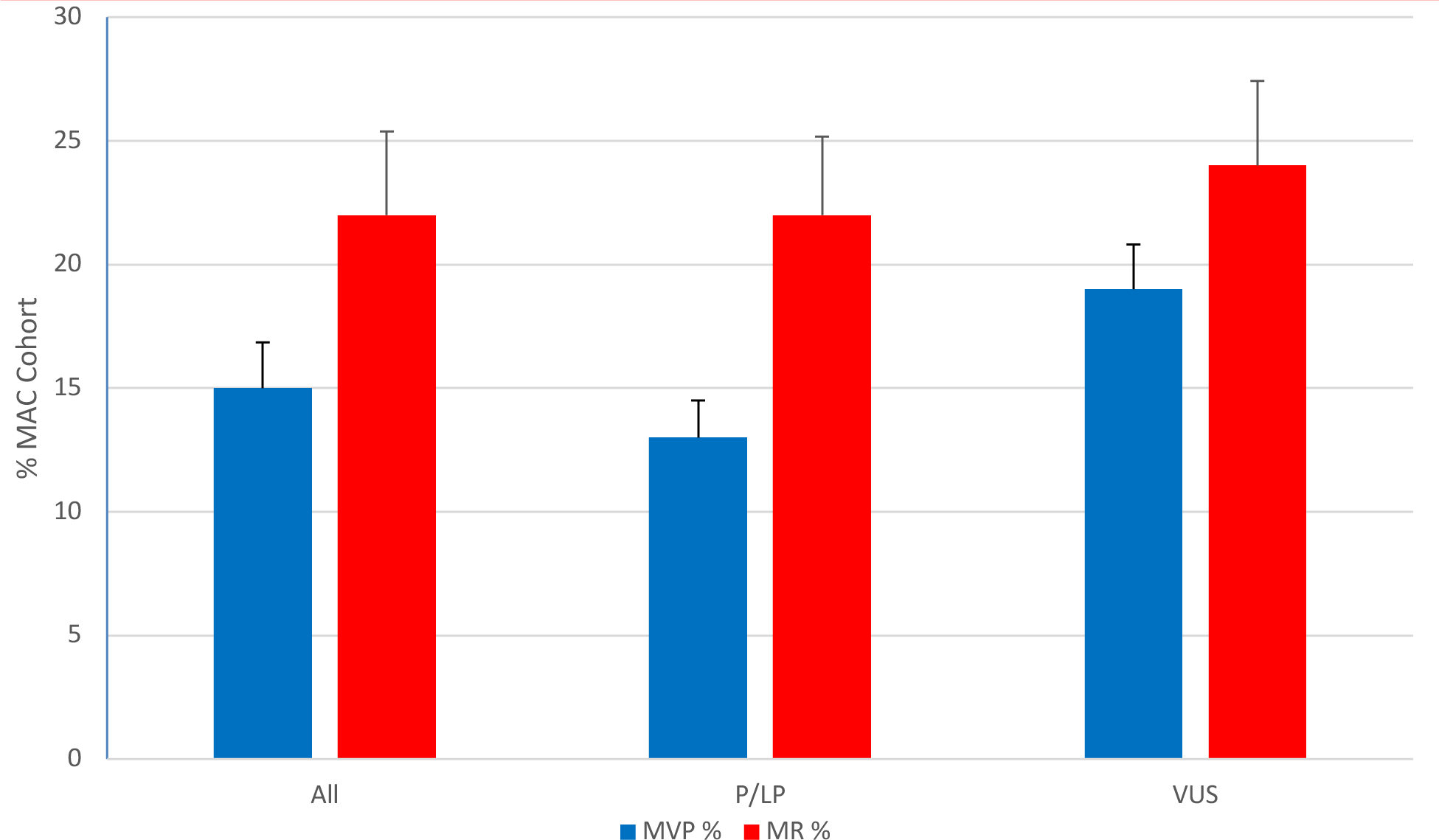
Prevalence of mitral valve prolapse and mitral regurgitation in MAC by variant classification. All: all variants, P/LP: pathogenic and likely pathogenic variants, VUS: variants of uncertain significance, MVP: Any degree of mitral valve prolapse, MR: mild, moderate, or severe mitral regurgitation.

**Figure S2.**
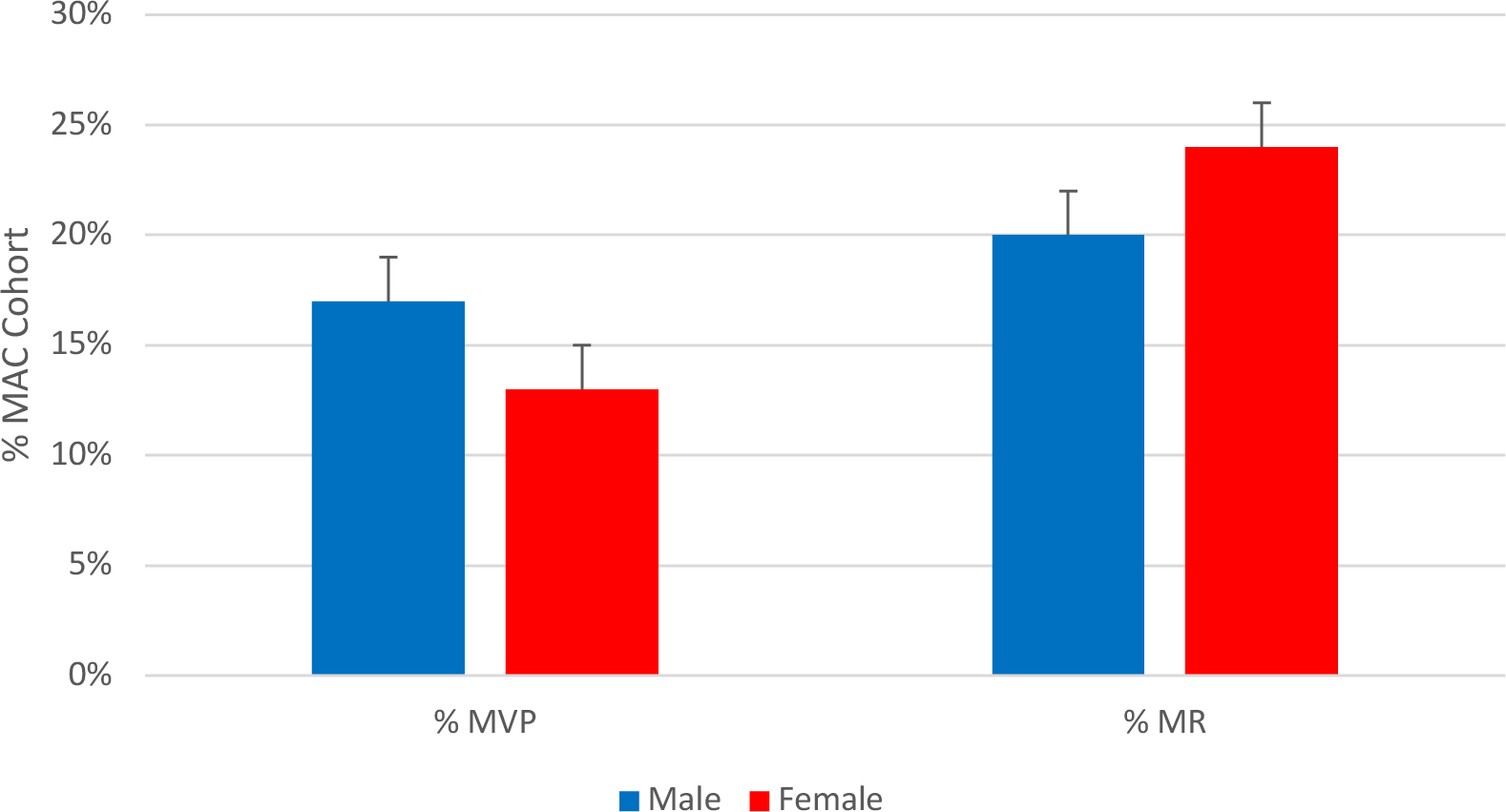
Prevalence of mitral valve prolapse and mitral regurgitation in MAC by sex. Mild, moderate, or severe mitral regurgitation was included in the analysis.

**Figure S3.**
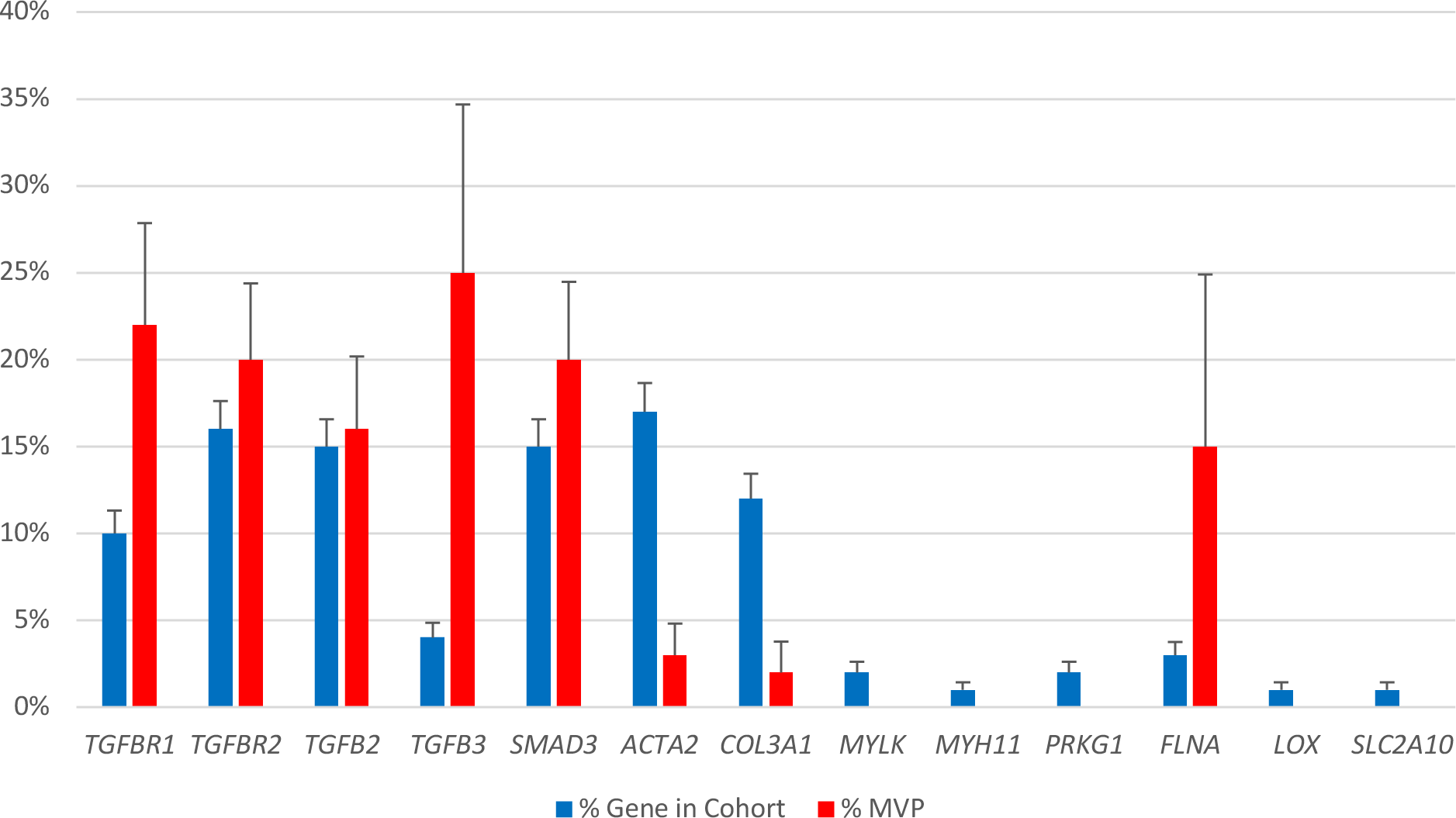
Prevalence of mitral valve prolapse and proportion of MAC participants with PV in each HTAD gene. MVP: any degree of mitral valve prolapse, %: percent cases. The prevalence of MVP was significantly enriched in MAC participants with PV in *TGFBR1* and *TGFB3*. *MFAP5, SMAD2,* and *SMAD6* were excluded due to <5 total cases.

**Figure S4.**
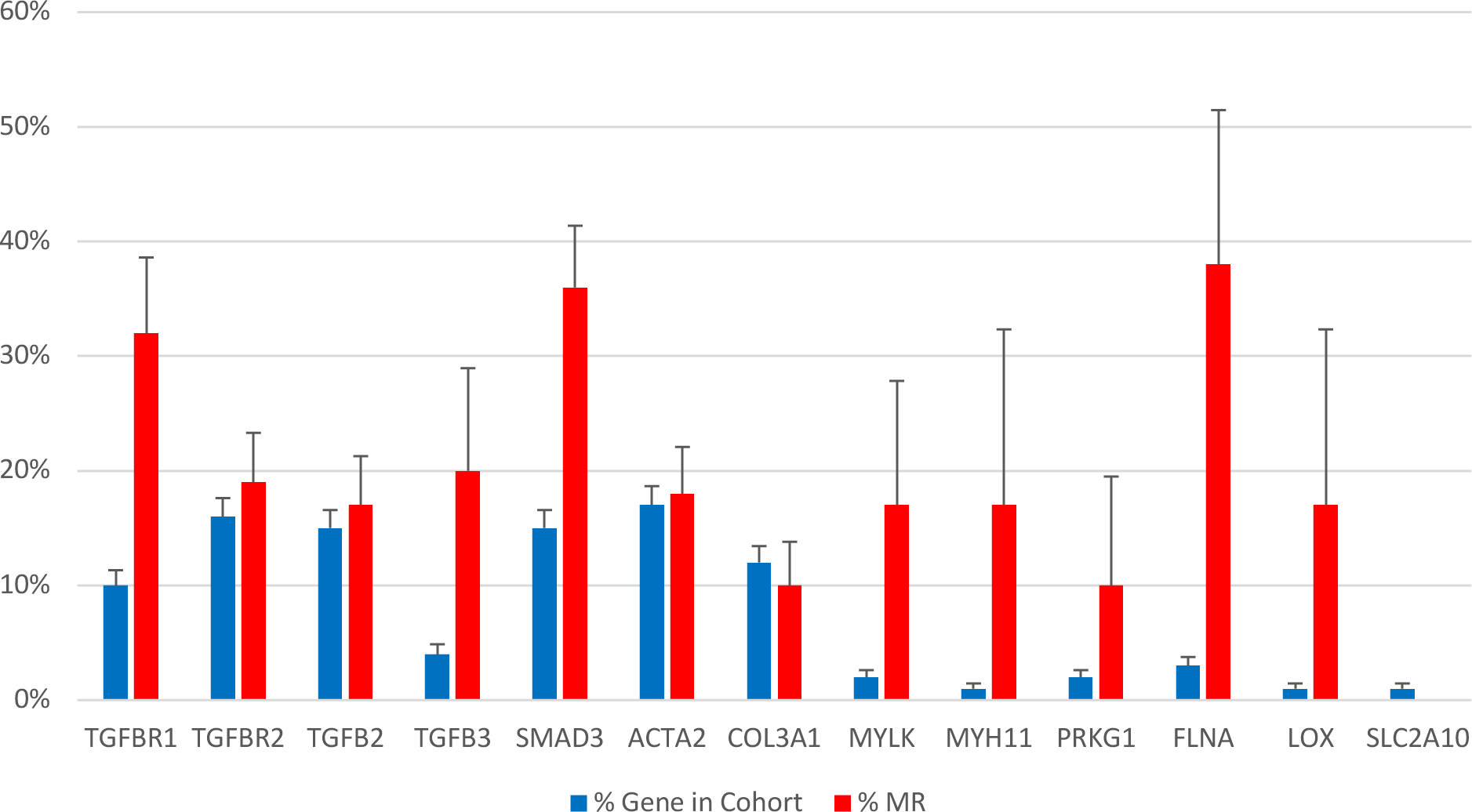
Prevalence of mitral regurgitation and proportion of MAC participants with PV in each HTAD gene. MR: mild, moderate, or severe mitral regurgitation. The prevalence of MR was significantly greater than expected for *TGFBR1*. *MFAP5, SMAD2,* and *SMAD6* were excluded due to <5 total cases.

**Table S1.**
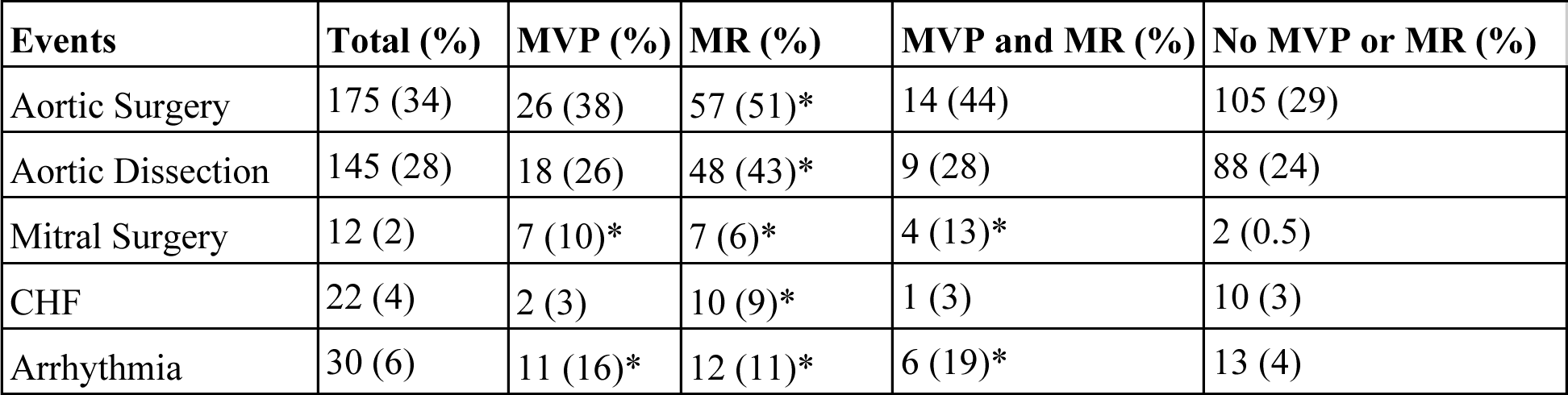
Cardiac and aortic events stratified by mitral phenotypes. MVP: any degree of mitral valve prolapse, MR: mild, moderate, or severe mitral regurgitation, CHF: congestive heart failure. Asterisk: *P*<0.05 vs. ‘No MVP or MR’ group.

**Table S2.**
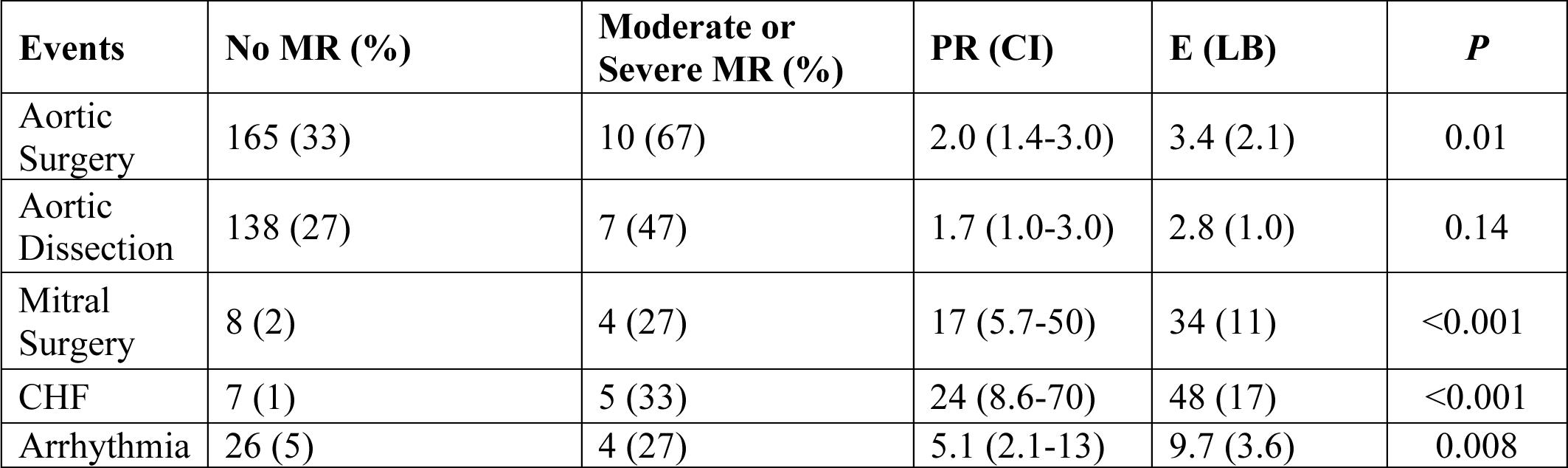
Cardiac and aortic events of MAC participants with moderate or severe MR. MR: moderate or severe mitral regurgitation, CHF: congestive heart failure, PR: prevalence ratio, E: E-value, CI: 95% confidence interval, LB: Lower bound. Percentages are in parentheses. Two-tailed *P*-values were calculated using Fisher’s exact test.

**Table S3.**
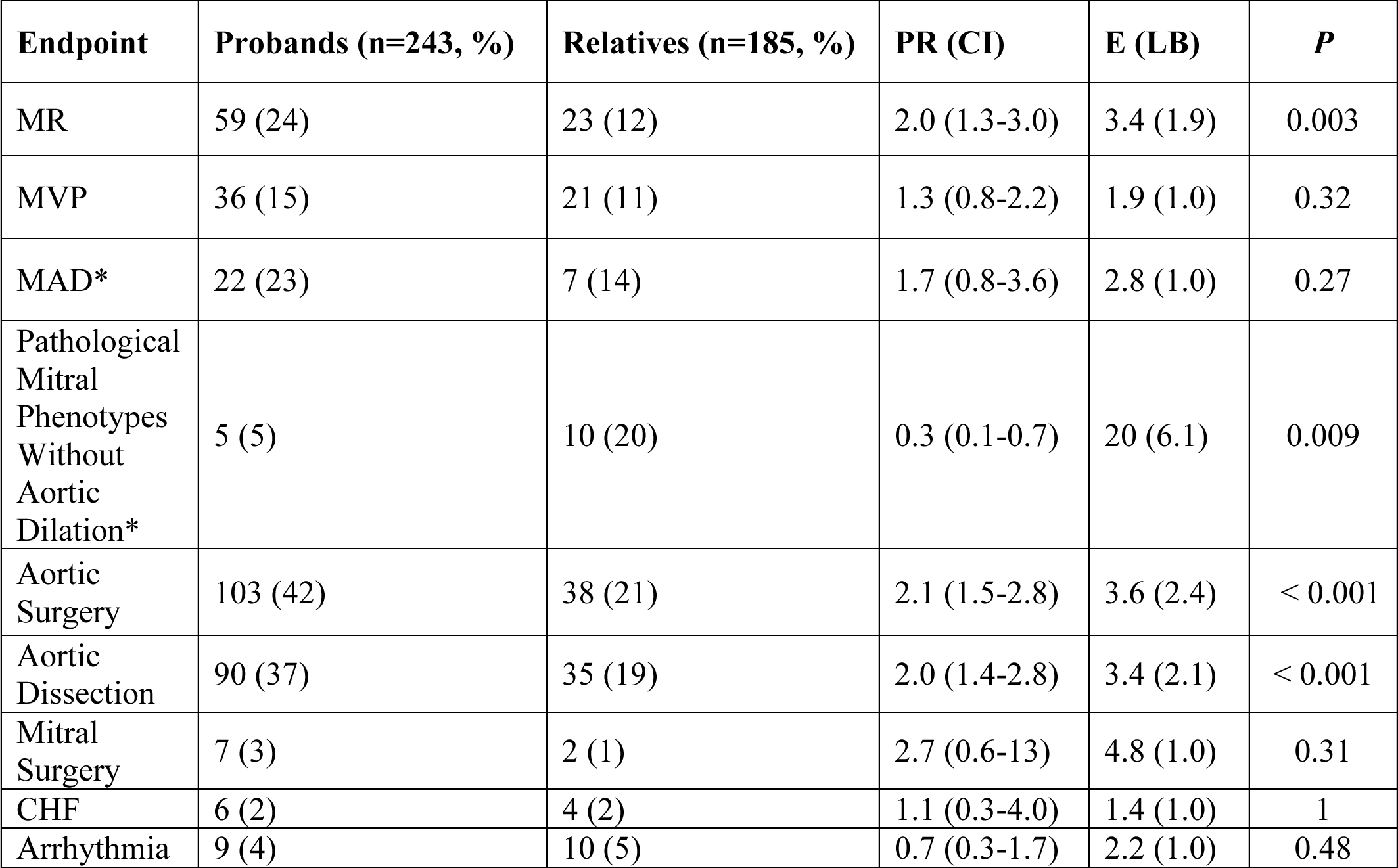
Characteristics of MAC participants by proband status. MR: mild, moderate, or severe mitral regurgitation, MVP: any degree of mitral valve prolapse, MAD: mitral annular disjunction, CHF: congestive heart failure, PR: prevalence ratio, E: E-value, CI: 95% confidence interval, LB: Lower bound. *Analysis was confined to MAC probands (n = 95) and relatives (n = 50) with available images. Aortic dilation: Z-score > 3, Pathological mitral phenotypes: >mild MR, ≥ 10 mm MAD, or MAD with MVP. MR, aortic surgery, and aortic dissection were more prevalent in probands, while predominant mitral phenotypes were more prevalent in relatives.

**Table S4.**
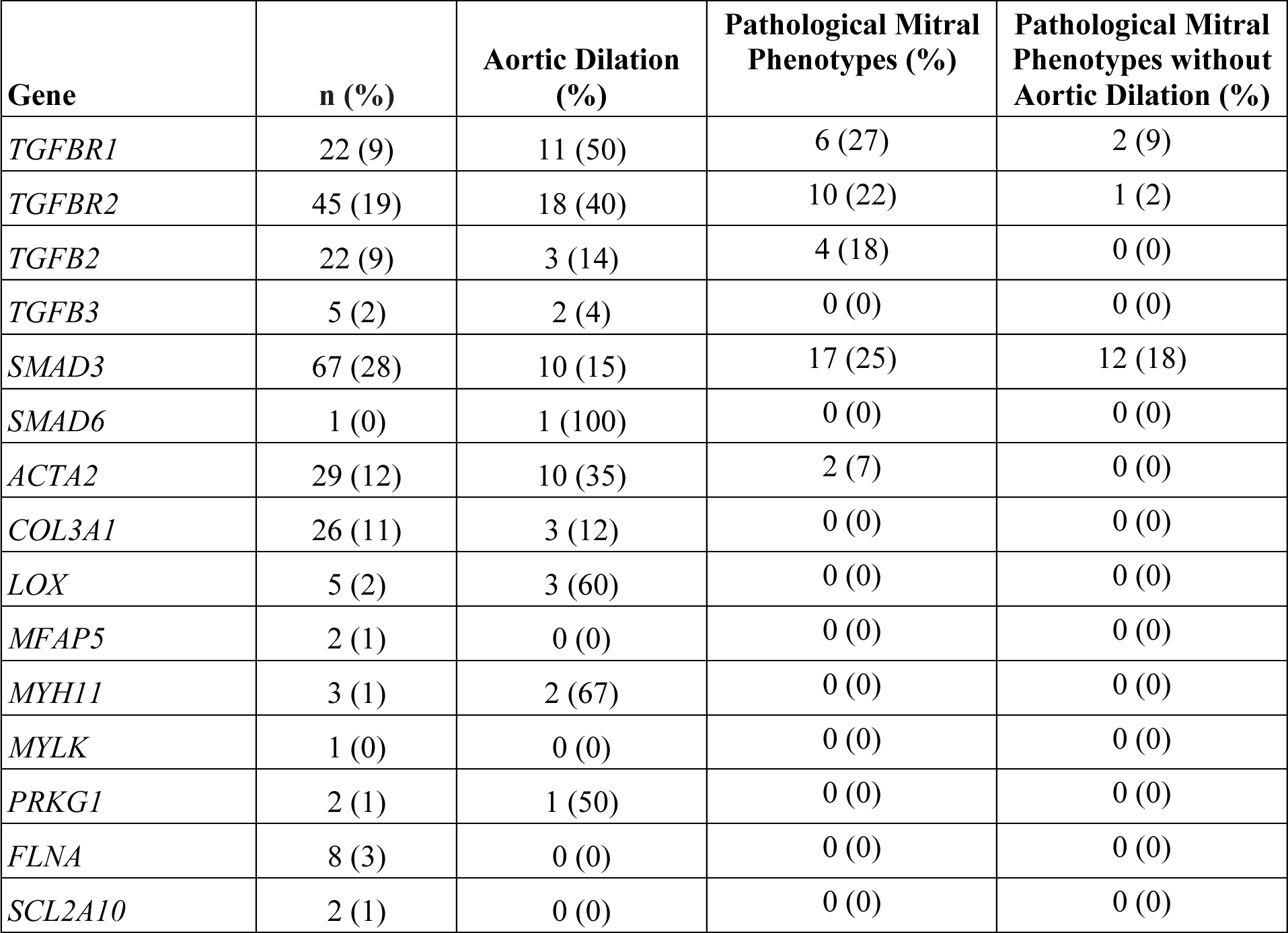
Proportion of MAC participants with pathological mitral phenotypes and/or aortic dilation by HTAD gene. Aortic dilation: Z-score > 3, Pathological mitral phenotypes: >mild MR, ≥ 10 mm MAD, or MAD with MVP. Analysis was limited to the subset of MAC participants with available images (n = 240).

**Figure S5.**
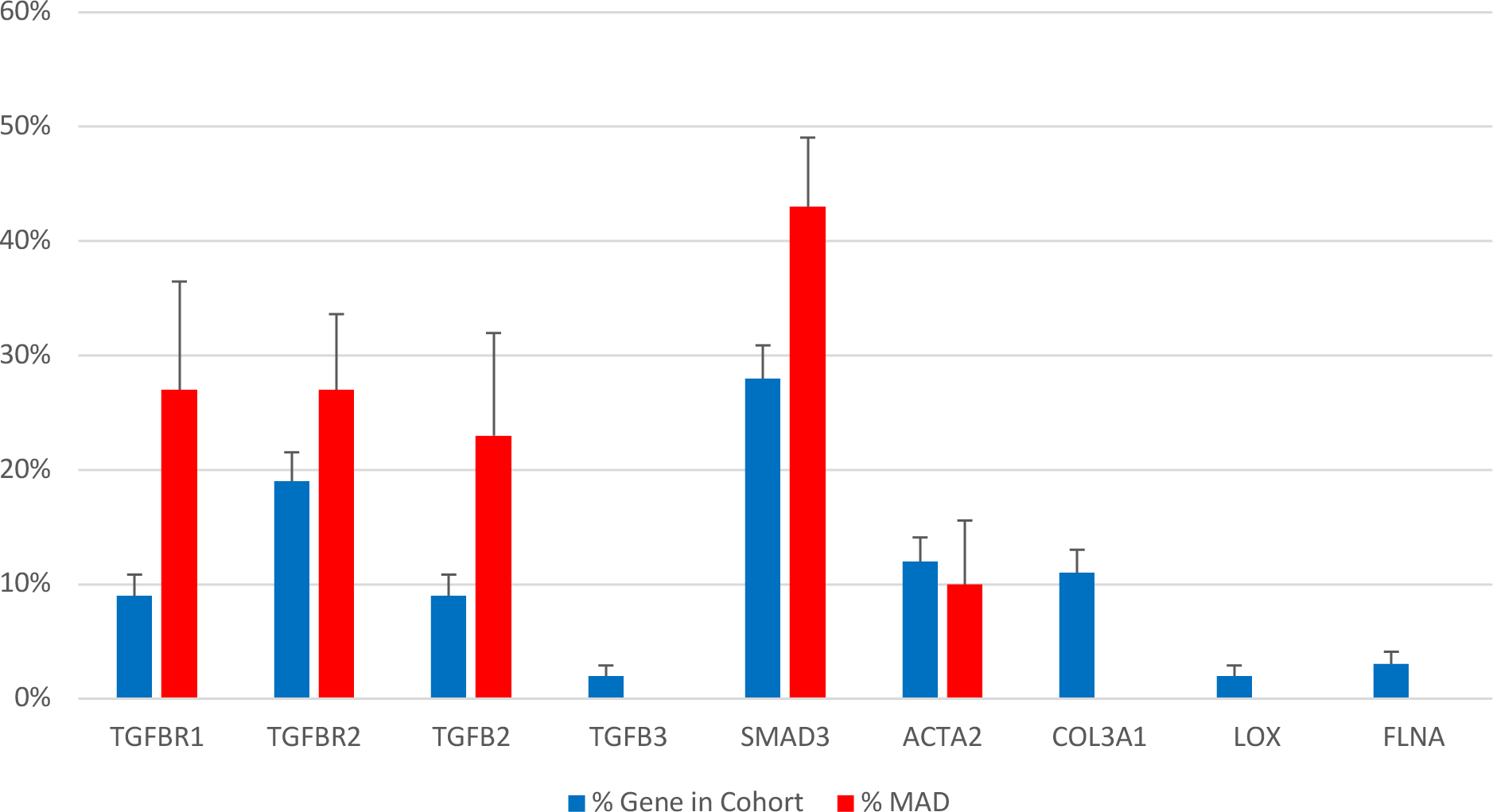
Prevalence of MAD and proportion of MAC participants with PV in each HTAD gene. MAD: mitral annular disjunction. The prevalence of MAD was significantly enriched in MAC participants with PV in *TGFBR1*, *TGFB2*, and *SMAD3*. *MFAP5*, *MYH11*, *MYLK*, *PRKG1*, *SMAD6*, and *SLC2A10* were excluded due to < 5 cases. Limited to the subset of MAC participants with available images (n=240).

**Figure S6.**
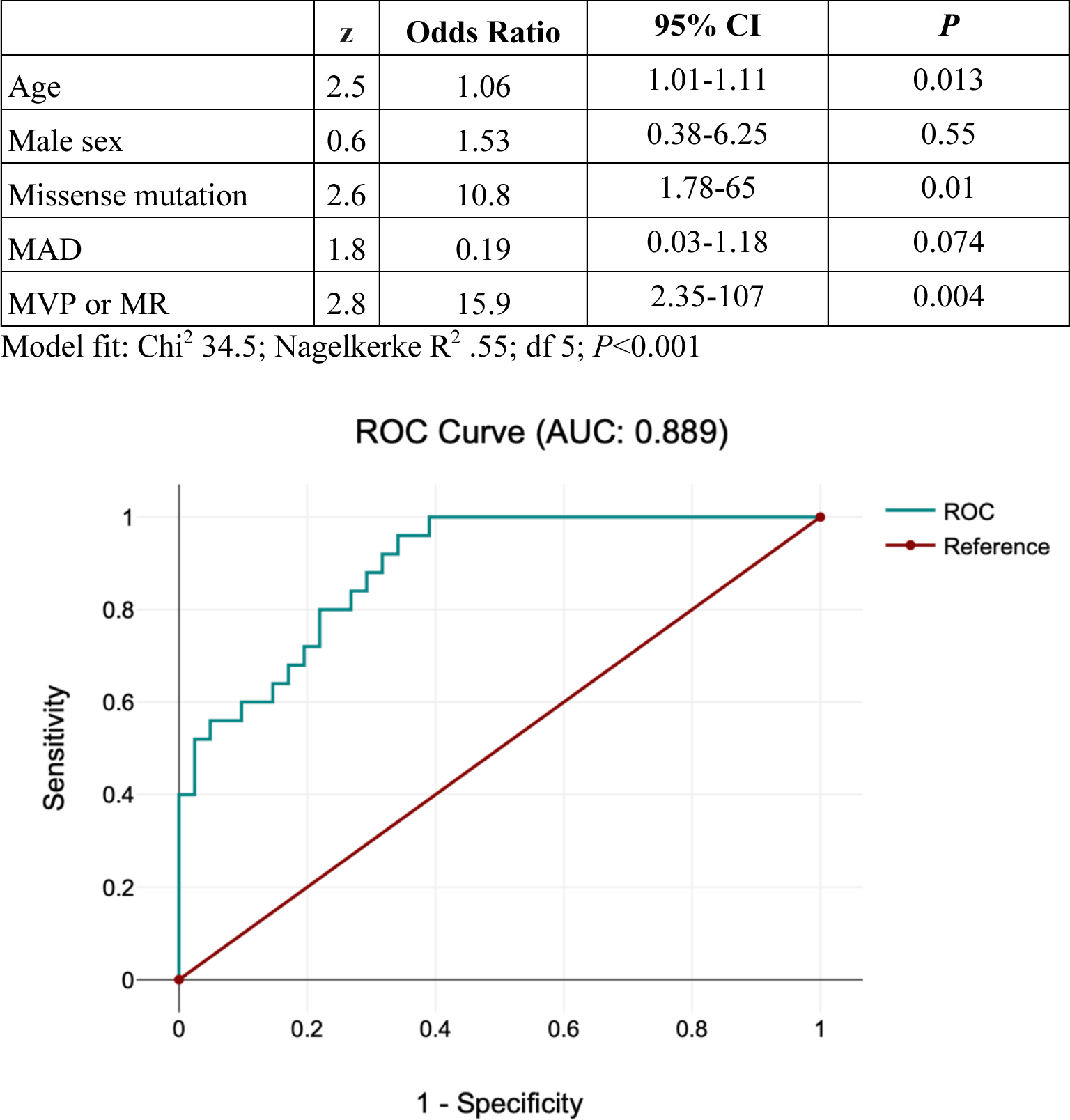
Logistic regression of composite clinical events (aortic surgery, mitral surgery, aortic dissection, arrhythmia, CHF) for MAC participants with *SMAD3* PV. MVP: any degree of mitral valve prolapse, MR: mild, moderate, or severe mitral regurgitation.

